# Anxiety, worry and perceived stress in the world due to the COVID-19 pandemic, March 2020. Preliminary results

**DOI:** 10.1101/2020.04.03.20043992

**Authors:** Rosario Sinta Gamonal Limcaoco, Enrique Montero Mateos, Juan Matías Fernández, Carlos Roncero

## Abstract

**Introduction:** Since the beginning of the outbreak in China, ending 2019, the Novel Coronavirus (COVID-19) has spread subsequently to the rest of the world causing an on-going pandemic. The World Health Organisation (WHO) declared COVID-19 “a public health emergency of international concern.” Taking into consideration the lockdown and quarantine situation, a research team of doctors from the Hospital of Salamanca, decided to do an evaluation of the current emotional state on the general population with a web-based survey in English and in Spanish, which was considered a useful and fast method that could help us determine how people perceived stress and worry due to the COVID-19.

**Methods:** The survey included a 22 items, gathering information in 3 sections: Sociodemographic data, the Perceived Stress Scale (PSS-10) by Cohen and additional queries that assessed the current worry and change of behaviours due to this pandemic.

**Results:** The survey received 1091 respondents from 41 countries, from March 17 to the 1^st^ of April, 2020. The mean age of the respondents was 43.1 (14.2) years old, and more than two thirds were women. 21.1% were health personnel. The mean of the PSS-10 score was 17.4 (6.4). Significantly higher scores were observed among women, youth, students, and among those who expressed concern and those who perceived increased susceptibility to the COVID-19. In contrast, no significant differences were observed between the health professionals and the general population. A weak correlation was observed between mean relative volume RSV of the last 28 days and the number of cases reported (rho = 0.31, p <0.001) and deaths (rho = 0.28, p <0.001).

**Discussion:** With these results the researchers describe an increase of affective symptoms due to the COVID-19. This pandemic is raising the anxiety levels. The findings of the study show the affective and cognitive alterations people are going through. This survey is the first attempt to measure the psychological consequences this pandemic is having, in order to be able to provide the support to confront this global issue, addressing the mental health care that will be needed.

## Introduction

Towards the end of 2019, the WHO China Country Office was informed of cases of pneumonia with an unknown aetiology detected in Wuhan City, Hubei Province of China. Further on, after trying to point out the cause of the outbreak, Chinese authorities identified a new type of coronavirus on January 7, called SARS-CoV-2. On January 12, 2020, China shared the genetic sequence of the novel virus. Since the beginning of the outbreak, what is now known as the Novel Coronavirus (COVID-19) has spread subsequently to the rest of the world causing this particular type of respiratory disease to be a pandemic. COVID-19 can cause anything from mild respiratory problems to pneumonia or death. Being men and older people the ones who mostly suffer the severity of this infectious desease. As the on-going pandemic continues to develop, World Health Organisation (WHO) declared COVID-19 “a public health emergency of international concern.” Approximately 60% of the global COVID-19 cases are currently outside China. At the time the survey was sent (March 17, 2020) the global situation registered by the WHO was: confirmed cases reported 207,860. Deaths 8,657 (Most of them in China and Europe-Italy and Spain mostly). Countries and territories with positive cases: 166. There is a very high rate of reported cases in health professionals.

The authors of this article, anticipated that the outbreak of COVID-19 would be stressful for people. Taylor et al. 2010, described the high levels of reliance on the support of others to get through tough times, for example, the vulnerability that may be felt by this potential global health threat which is requiring the use of physical control measures, such as social distancing, home quarantine, and school and work closures, all resulting in disruption to social support networks at a time when they may be needed most. Brooks et al. 2020, informs that the psychological impact of quarantine is wide-ranging, substantial, and can be long lasting. This change of circumstances and raise of stressors has to make psychiatrists be attentive to possible relapses in patients with mental health problems. Taking into consideration the lockdown and quarantine situation, the researchers decided to do evaluate the current state on the general population with a web-based survey that was considered at this moment a useful and fast method that can help determine how people perceived stress and worry due to the COVID-19. Feizi et al. 2012, points out that psychological stresses are also associated with huge increase of mortality in general population, if the quarantine experience is negative, it suggests there can be longterm consequences that affect not just the people quarantined but also the healthcare system as referred to the Brooks et al. 2020, study. In order to determine the current and future support people will need to confront this situation.

This study aims to observe various points. First, it aims to find out how this pandemic is progressing and how it’s producing changes in the affective state of the general population. Next, it is expected, that the higher risk groups should score higher in the stress scale, such as older people and health professionals due to the overwhelming situation they are living in hospitals around the world. This survey is the first attempt to measure the psychological consequences this pandemic is having. The results of the study will help determine the mental health care that might be needed.

## Methods

Counting that the stress perception founds on cultural and social aspects that can vary from one country to another. On March 17, 2020 we sent a web-based survey with an English or Spanish version. Both versions were chosen for being the native language in many countries, as well as a good option for those countries who don’t natively speak this languages. This study was endorsed by the member in charge of the Research Ethics Committee, of the University of Salamanca Health Care Complex. The survey was sent to mainly Madrid and Manila, and other cities in the world. At that time both cities had already a law enforced lockdown by their governments. The recruiting of the rest of participants was on forums and social networks.

Those who received the survey and were interested in participating answered the questionnaire freely, knowing that the survey’s participation was voluntary and all data is anonymous. The questionnaire had 22 items, that gathered information in 3 sections:

1. Sociodemographic data, including age, gender, nationality, and employment status, as well as the current city they were at at the time of answering the questionnaire and a dichotomous question to specify if they were a health professional or not.
2. Supplementing this, they answered the Perceived Stress Scale (PSS-10; Cohen et al., 1988) with 10 items, which was chosen for it being a widely used psychological instrument that measures the degree to how circumstances in one’s life are detected as stressful. Designed to determine how unpredictable, uncontrollable, and overloaded respondents find their lives, which we found ideal for this current situation. Baik et al. 2017, show in their study the scores on the PSS-10 were significantly correlated in the expected directions with scores compared in these scales: Generalized Anxiety Disorder-7 (GAD-7) and Patient Health Questionnaire-9 (PHQ-9), in both English and Spanish language preference groups.
3. In addition, based on the study of the influenza A/H1N1 pandemic of Liao et al. 2014, they answered a question per item to estimate (all related to COVID-10): the anticipated worry (a prospective measure), experienced worry (a retrospective measure), current worry (a current measure), perceived absolute susceptibility (a prospective measure), perceived relative susceptibility (a prospective measure) and two other questions related to the altered/or not behaviour due to the COVID-19 related to the CDC recommendations: avoiding crowded places and hand cleaning.

Furthermore, some epidemiological data was obtained from different sources (countries departments of health, newspaper) to explore the possible association of them with the PSS-10 score: days since the first case reported, number of cases and death reported the day before in the country. Furthermore, in order to assess the social impact of the disease and its possible correlation with the perceived stress, mean relative volume (RSV) of the 7 and 14 days of each country was obtained from Google Trends, using the terms “coronavirus” and “COVID”.

Dichotomic variables were analysed using chi-square test, U Mann-Whitney (or Kruskal Wallis when required) was used to assess the differences between non-parametric variables. Spearman’s rho was used to measure the correlation between viables. Finally, multiple linear regression with backward elimination was conducted to identify independent factors that determine the PSS-10. Statistical analysis was performed using SPSS package v 20.0.

## Data analysis and results

The survey received 1091 respondents from 41 countries(figure 1), from the 17th to the 1^st^ of April, 2020. The majority of participants were from the Philippines (43%) and Spain (30%) and Colombia (11%). In almost all the countries that responded, COVID-19 cases had already been declared before answering the survey (median 539 cases, 142-9191) and the majority of the responses came from countries with reported deaths (98%). The mean age of the respondents was 43.1 (14.2) years old, and more than two thirds were women. 72.6% of the respondents were part-time or full-time workers and 21.1% were health personnel (Table 1).

**Table 1.**

|  | Frequencies (%) |
| --- | --- |
| **Sex**<br>- Female<br>- Male | - 741 (67.9%)<br>- 350 (32.1%) |
| **Age**<br>- <30 yr<br>- 30-59 yr<br>- 60+ yr | - 244 (22.4%)<br>- 706 (64.7%)<br>- 139 (12.7%) |
| **Employment Status**<br>- Employed<br>- Unemployed<br>- Student<br>- Other | - 792 (72.6%)<br>- 68 (6.2%)<br>- 73 (6.9%)<br>- 156 (14.2%) |
| **Healthcare profesional**<br>- Yes<br>- No | - 227 (20.8%)<br>- 864 (79.2%) |
| **Region:**<br>- South Eastern Asia<br>- Southern Europe<br>- South America<br>- North America<br>- Western Europe<br>- Western Asia | - 478 (43.8%)<br>- 350 (32.1 %)<br>- 145 (13.3%)<br>- 39 (3.6%)<br>- 21 (1.9 %)<br>- 15 (1.4%) |
| **National quarantine**<br>- Yes<br>- No | - 974 (89.3%)<br>- 111 (10.2%) |
| **Anticipatory worry**<br>- Yes<br>- No | - 685 (62.8%)<br>- 406 (37.2%) |
| **Experienced worry**<br>- Yes<br>- No | - 823 (75.4%)<br>- 268 (24.6%) |
| **Current worry**<br>- 0-5<br>- 6-10 | - 180 (16.5%)<br>- 911 (83.5%) |
| **Perceived susceptibility to COVID**<br>- Likely<br>- Unlikely | - 206 (18.9%)<br>- 885 (81.1%) |
| **Perceived relative susceptibility to COVID**<br>- Higher<br>- Lower | - 222 (20.3%)<br>- 869 (79.7%) |
| **Avoiding crowded places due to COVID**<br>- Yes<br>- No | - 573 (52.5%)<br>- 518 (47.5%) |
| **Hand Hygiene measures**<br>- Often or very often<br>- Less frequent | - 735 (67.4%)<br>- 356 (32.6%) |

**Figure 1.**
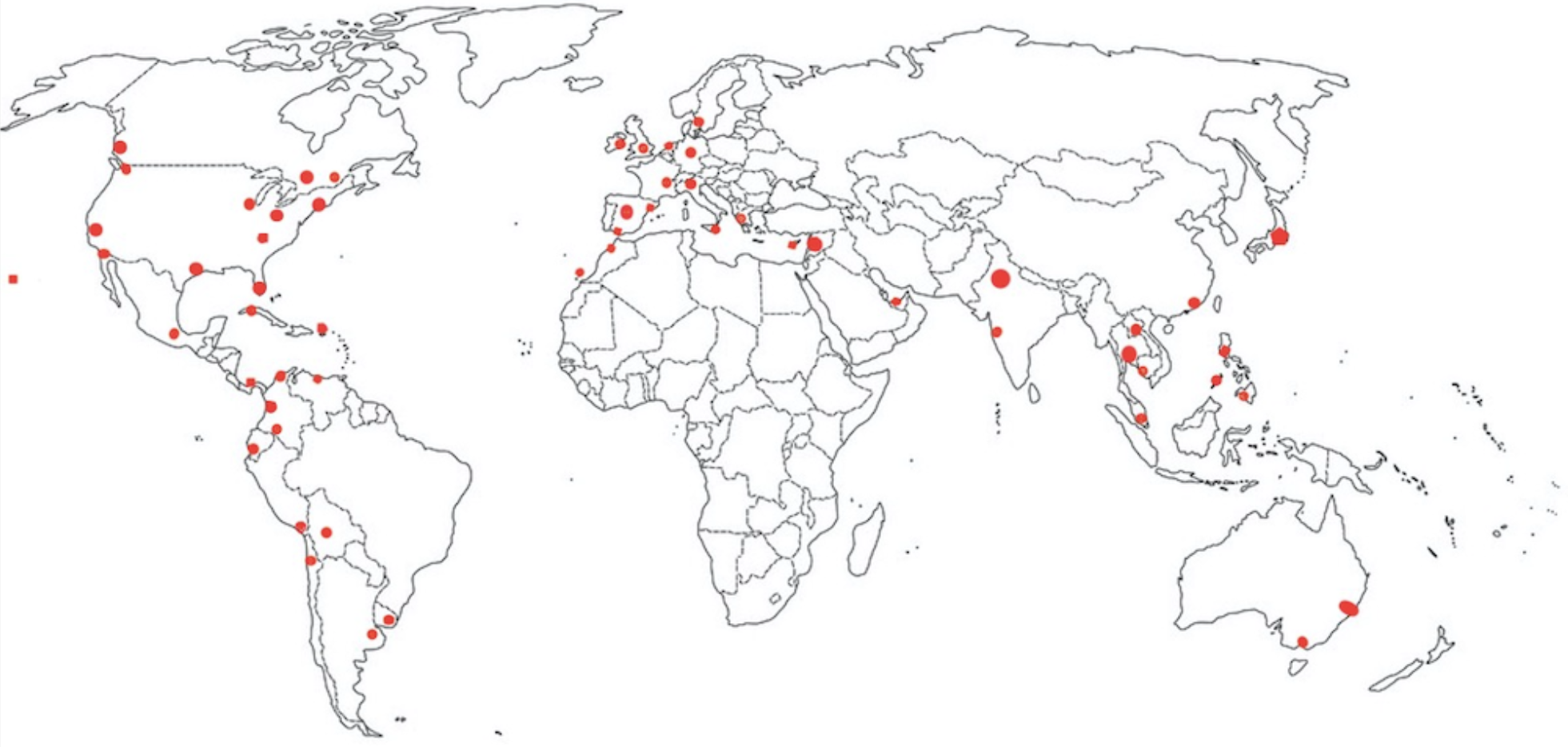

The mean of the PSS-10 score was 17.4 (6.4). Significantly higher scores were observed among women, youth, students, and among those who expressed concern about becoming infected with COVID-19 and those who perceived increased susceptibility to the coronavirus (Table 2).

**Table 2.**
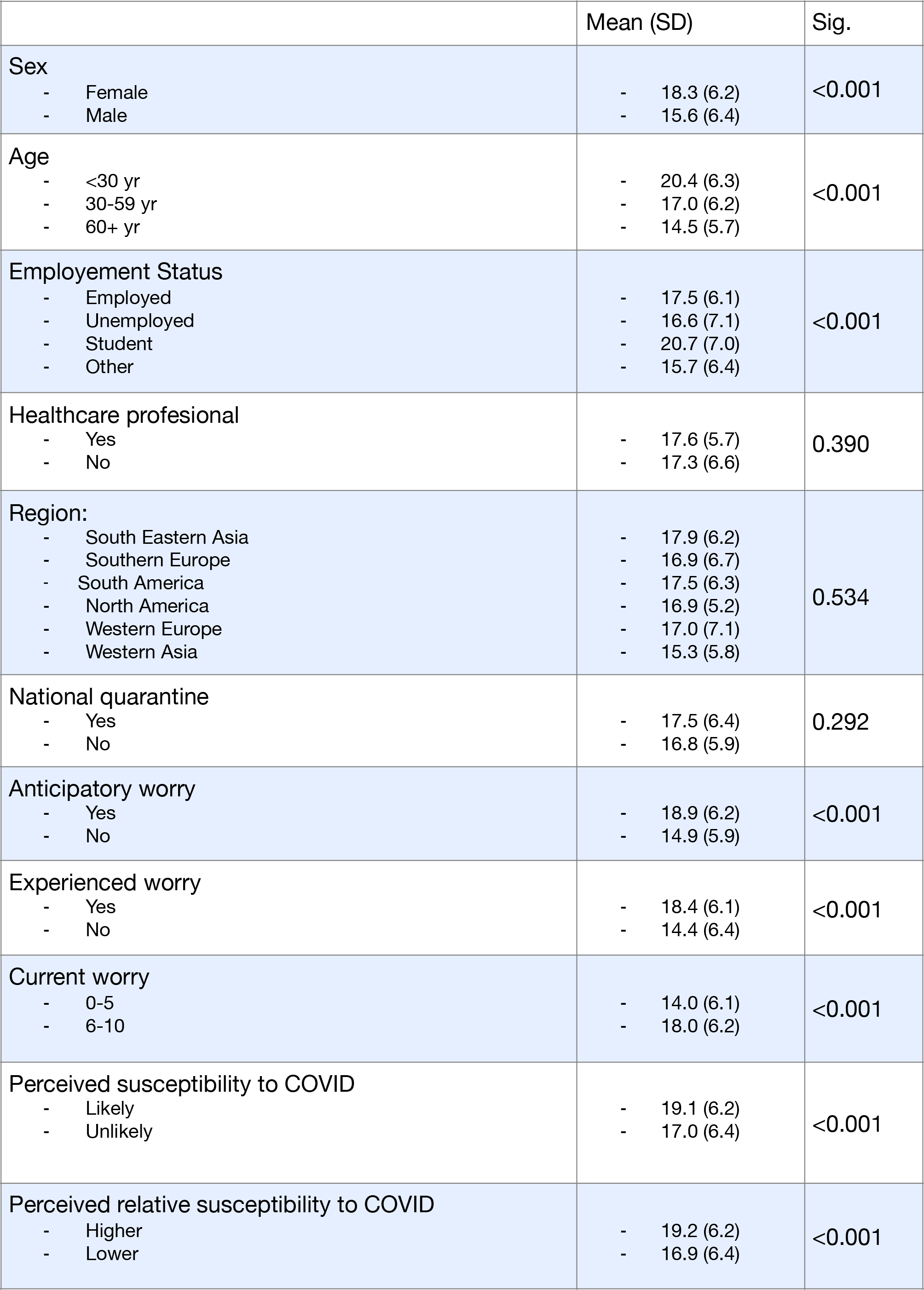
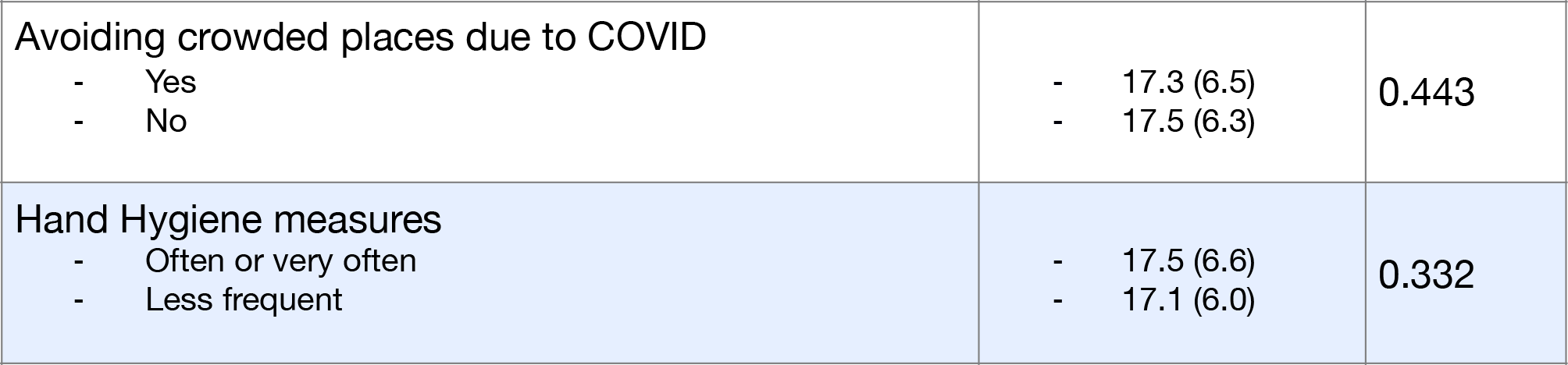

In contrast, no significant differences were observed between the health professionals and the general population, or between those from quarantined countries and those who did not have quarantine in their country at that moment.

Avoiding crowded places was significantly more frequent among those with the anticipated worry (56.9.7% vs 45.1%, p<0.001), experienced worry (56.6% vs 39.9%, p<0.001) and current worry (54.3% vs 43.3%, p=0.007), showing no significant association with susceptibility questions. Hand hygiene was significantly more frequent among those with: anticipated worry (71.5% vs 60.3%, p<0.001), experienced worry (69.7% vs 60.1%, p=0.003), current worry (70.7% vs 50.6%, p<0.001) and perceived absolute susceptibility (74.3% vs 65.6%, p=0.013). None of those behaviours were statistically associated with higher PSS-10 scores.

In the linear regression, the items that perform as variable predictors of the PSS-10 were: constant (B 17,6, p<0.001), age (B -0.14, p<0.001), female (B 1.7, p=0.001) and the sum of responses about worry (B 2.1, p<0.001). Those variables explained 25% of the variance (R^2^=0.25).

An exploratory analysis was performed to assess the possible relationship of the PSS-10 score with the epidemiological impact in the country and public interest in the disease. A weak correlation was observed between the mean RSV of the last 28 days and the number of cases reported (rho = 0.31, p <0.001) and deaths (rho = 0.28, p <0.001), but none of them showed a significant correlation with the PSS-10 score.

## Discussion

These results describe heightened affectivity symptoms due to the COVID-19. This pandemic is raising the anxiety levels. Overall the sample showed a PSS-10 score higher than that reported in the general population (<13 according to Cohen et al. 1988) and close to the values reported by Chua et al. 2004, during the SARS outbreak in 2003, which placed it at 18.5. Furthermore, in our cohort those who reported concern or susceptibility to COVID-19 were those with higher scores.

Although the very nature of natural and health disasters is one synonymous with unpredictability, the results of the study show the affective and cognitive alterations people are going through. Mackay et al. 1978, described mood adjective checklists as a popular method of gathering data about an individual’s phenomenological perception of the behavioural and cognitive components of his reaction to different situations.

It could seem contradictory to find that older respondents showed lower levels of anxiety and worry even though this age group is reported as a high risk group. Pneumonia causes a more severe condition and even higher mortality among the elderly, as studied in Yang et al. We find a negative correlation between age and the score given on the scale. A statistically significant difference is observed between the median score between those over 60 years of age and those under that age. Remor E 2006, also shows that the stress scores had a tendency to decrease with age using the PSS, suggesting that perceptions of stress tend to decline as age increases, that result must be interpreted with care, as the correlation was very low. In this study, the findings are similar, older people present lower anxiety levels, even though they are the population with higher risk of presenting worse progress in this disease. Carstensen et al. 2006, offered reasons for this decline of stress with age, from the selectivity of positive aspects to reduced physical reactivity due to physical and health limitations. Frazier et al. 2019, emphasises three key components to explain the changes in integration for social decisions in ageing: theory of mind, emotion regulation, and memory for past experience.

The reports from China, Italy and Spain at present address a high rate of infections in the health professionals. They are the ones who attend to all cases and are high risk group due to the continuous exposure to patients with COVID-19. Although, these findings reflect that the heath personnel get the same stress levels/ PSS-10 score, than the general population, just as described by Chua et al. 2004, with SARS in 2003. With a similar score McAlonan et al. 2007, reports 17 (5.7), describes that health professionals at the moment of the outbreak, do not present higher levels of perceived stress. However they maintain high stress levels for a longer time turning it into chronic stress. A possible explanation to this, is that this professionals are more accustomed to managing higher stress levels because of the nature of their jobs. They are used to encountering critical health situations. Furthermore, the survey was carried out at the beginning of the infectious outbreak in many countries, the distress in this personnel was just beginning and could be minimised at that moment. To examine whether the stress level grows in time, alongside the progression of the pandemic would be interesting. Many countries have already declared this as a health emergency, taking measures in order to minimise transmissibility, morbidity and mortality; minimising the burden on health care systems.

The results of women presenting higher scores of anxiety, was already described by the initial author Cohen et al. 2012, and Remor E 2006, regarding PSS and PSS-10, women received significantly higher perceived stress scores than men. The reasons for this finding may be related to sex differences in coping with stress. The findings in this paper point out higher levels of anxiety and worry in women. Numerous studies, Uhlenhuth et al. 1973, show how women report higher intensity of the symptoms than men, and Kendler et al. 2001, displays gender and sensitivity to the depressogenic effect of stressful life events, where women reported higher stress rates. The study of Dalgard et al. 2006, explains why with a more affiliative style, and a stronger involvement in household and family matters, women are more exposed to problems in the social network. Based on this, women are more likely than men to report events in the social network, as it shows the contribution in each gender to this survey.

The main finding in the Liao et al. 2014, study is that affective measures of risk perception generally had stronger associations with reported modification of health protective behaviours. The behaviours examined in this survey to reduce the exposure to the infection were: the higher concern in avoiding crowded places and performing hand sanitising conducts almost all the time. No significant correlation was found between the reported cases and deaths of COVID in the country at the time of the survey and the PSS-10 score. Additionally, results did not identify significant differences in stress levels between countries that are currently in quarantine and those that are not, although this finding is limited as the sample of non-quarantined countries is small. As previously described in other infectious outbreaks, Bragazzi et al. 2017, at country level there was a correlation between digital interest toward coronavirus and epidemiological data (total cases and death reported), which was not found in this study with the PSS-10. Nevertheless, further studies will be required to explore the possible association.

One of the future objectives is to assist health workers and the general public in managing emotional stress and related personal, professional and family issues during the COVID-19 pandemic. While there is currently no recommended treatment, vaccine, or antiviral medication for COVID-19, pharmaceutical companies are racing for solutions. Our response to mitigate the affective and cognitive consequences on the quarantine can be based on the Brooks et al. 2020, recommendations: giving people as much information as possible, providing adequate supplies, reducing the boredom and improving communication.

### Limitations

The results of this study should be interpreted within the context of study limitations. Methodological issues limit the validity of the research results. In addition the target population is not specific, our study comprised responses from 41 countries, the response from individual countries was not uniform, three countries were mostly represented as well as the demographic bias in the sample, the particular greater proportion of women, which is related as mentioned before with the greater interest for participation in studies of this nature. Furthermore, it is important to point out that future studies should also address the different demographics of the health-care system in each country.

Although the items of this tests based on self-observation also bear a certain degree of ambiguity, for example, by using the term “often,” that can be interpreted in various ways, projective techniques, in general, are providing a much wider freedom of response and consecutively provoking a much wider response variety in nature and number going together with a complex procedure of scoring. Therefore, they are much more vulnerable by interpreter’s scope of accuracy.

Based on this description, the researcher propose a follow up and extension of this study to be able to reach more countries and know the mental health impact this pandemic is having. The changes that are assessed can lead to affective and cognitive problems, which should be able to be assisted by health professionals. Creating health interventions and preparing to aid mental health issues due to this pandemic is the main objective.

## Data Availability

The data in this manuscript is referred and available in the manuscript itself.

